# High mortality and complications in patients admitted with Takotsubo cardiomyopathy without improvement in outcomes over the years

**DOI:** 10.1101/2024.06.19.24309204

**Authors:** Mohammad Reza Movahed, Elimira Javanmardi, Mehrtash Hashemzadeh

## Abstract

**Introduction:** Takotsubo cardiomyopathy continues to be a major cause of mortality and morbidity. The goal of this study was to evaluate the outcome data of patients with Takotsubo cardiomyopathy using a large inpatient database.

**Methods:** We used the Nationwide inpatient sample (NIS) database for our study. We evaluated trends, mortality, and complications of patients admitted with Takotsubo cardiomyopathy from available 2016-2020 years in adults over the age of 18.

**Results:** A total of 199,890 patients with Takotsubo were found in our database with 83% being female and higher prevalence with age, Caucasian race, and highest income. Mortality was high at 6.5 % with no significant improvement over the years studied. Furthermore, major complications were substantial. Cardiogenic shock occurred in 6.6%, atrial fibrillation in 20.7%, Cardiac arrest in 3.4%, congestive heart failure in 35.9% and stroke in 5.3%.

**Conclusion:** Takotsubo cardiomyopathy is associated with high mortality and complications with no improvement in outcome over the 5 years study. Further improvement in care is needed to improve outcomes.

## Introduction

Takotsubo cardiomyopathy (TC) is a reversible left ventricle dysfunction and is triggered by emotional stress, predominantly in women, or physical stress, predominantly in men [1, 2]. This condition is known to be associated with gender and race disparities and can lead to significant in-hospital mortality and morbidity [3, 4]. Despite the recovery in a later time, little is known about the course and compilations of this condition over the recent years. The goal of this study was to use the largest database available to determine the incidence of TC and associated cardiovascular complications through the years 2016-2020 using the Nationwide inpatient sample (NIS).

## Methods

### Data source

The study data was derived from the Nationwide Inpatient Sample (NIS). The NIS database includes unweighted hospitalization information for about 7 million and weighted data information for about 35 million hospitalizations [5]. Review Board approval was not required since the NIS data are publicly available.

### Study population and outcomes

In this retrospective observational cohort study, all patients aged over 18 years from the 2016-2020 NIS database were included. Our target population was patients hospitalized with TC which was identified using ICD-10 code I51.81. The key outcomes of interest including myocardial rupture (I23.2, I23.3, I23.4, I23.5), cardiogenic shock (R57.0), atrial fibrillation(I48), cardiac arrest (I46), congestive heart failure (I52, I53, I54), and stroke (I60, I61, I62, I63) were also recorded. Patient demographics include age, gender, race, and hospital demographics including median household income, expected primary payer, hospital bed size, location and teaching status of hospital, hospital region, control of hospital along with average hospital length of stay (LOS), and average total charges were evaluated.

### Statistical Analysis

Patient demographic, clinical, and hospital characteristics are reported as percentages in the tables. Odds ratios and 95% confidence intervals are calculated for continuous variables and proportions, and 95% confidence intervals for categorical variables. Trend analysis over time was assessed using Chi-squared analysis for categorical outcomes. Multivariable logistic regression ascertained the odds of binary clinical outcomes relative to patient and hospital characteristics as well as the odds of clinical outcomes over time. Multivariable linear regression will estimate the mean differences of length of stay relative to patient and hospital characteristics. All analyses were conducted following the implementation of population discharge weights. All p-values are 2-sided and p<0.05 was considered statistically significant.

Data were analyzed using STATA 17 (Stata Corporation, College Station, TX).

## Results

### Baseline characteristics

In the investigation of the NIS database from 2016 to 2020, a weighted total of 199,890 patients who were hospitalized with Takotsubo cardiomyopathy were identified based on the ICD-10 codes. The annual incidence of TC patients did not show a consistent pattern but overall increased from 39,015 in 2016 to 41,290 in 2020.

The baseline characteristics are presented in Table 1. The mean age was 67.09±14.15 years, and 83% of cases were female. As shown in Figure 1, incidence increased from 0.19% to 0.21% for females and from 0.05 to 0.07 % of all hospitalizations for males, respectively, during the study years.

**Table 1.** Patient Characteristics (%) (N = 199,920)

| Table 1. Patient Characteristics (%) (N = 199,920) |  |
| --- | --- |
| Mean ±SD age (years) | 67.09±14.15 |
| Female | 83 (165120) |
| Male | 17 (34770) |
| **Race** |  |
| White | 77.76 (155475) |
| Black | 7.93 (15855) |
| Hispanic | 6.17 (12345) |
| Asian/Pac Isl | 2.05 (4105) |
| Native American | 0.61 (1235) |
| Others | 2.2 (4400) |

**Figure 1.**
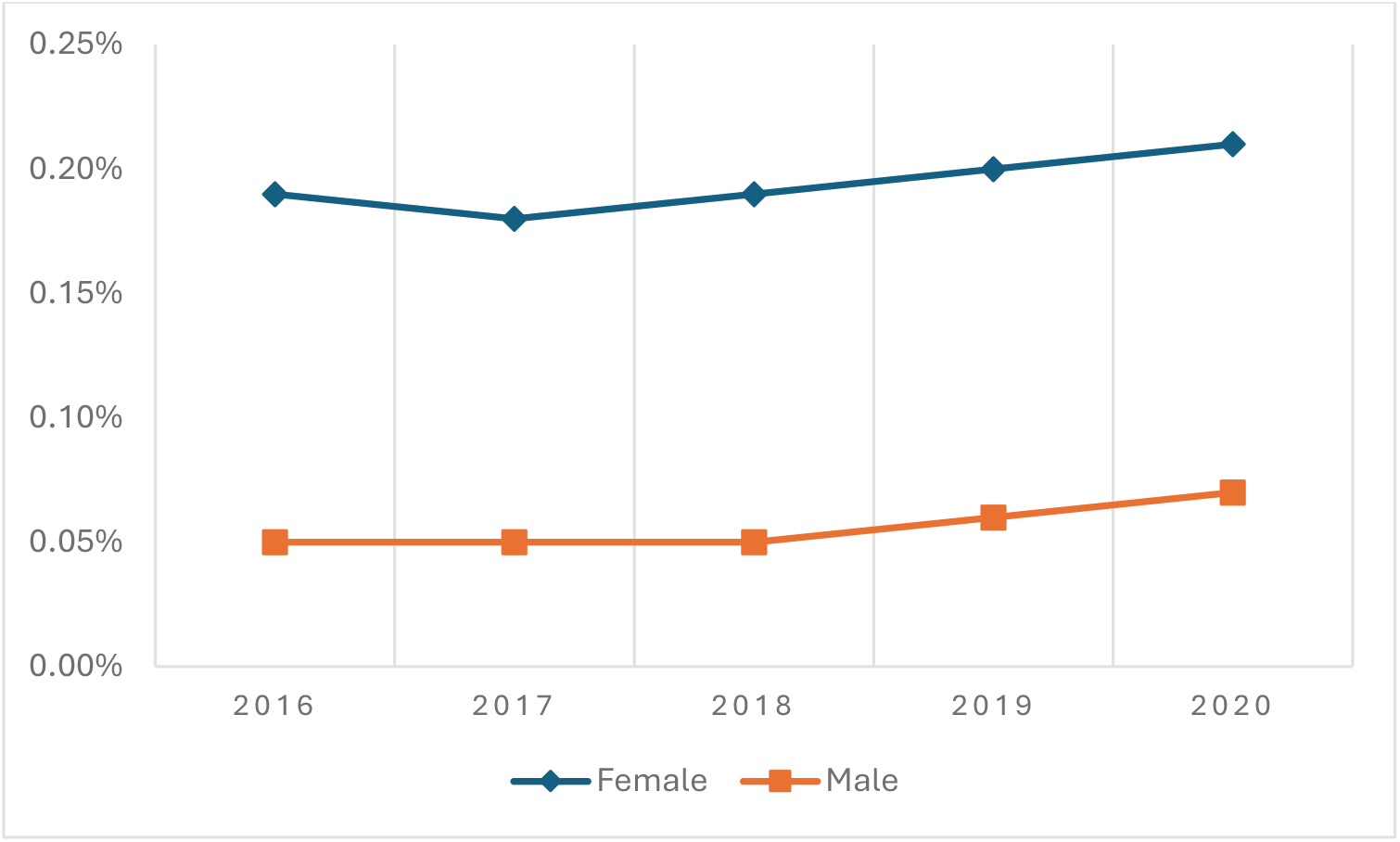
Takotsubo Cardiomyopathy Incidence by Gender.

The TC incidence increased across all age groups from 2016 to 2020 shown in Figure 2. The intra-group change was highest in 46-60 age group with 3% change, and lowest in the 18-30 and 31-45 age groups with 1 % change, respectively. Age groups older than 61 showed a 2% change from 2016 to 2020. The age group above 61 consistently had the highest incidence rates, from 0.19 % in 2016 to 0.21% in 2020, while the age group 18-30 had the lowest incidence rates, from 0.01% in 2016 to 0.02% in 2020. Importantly, there was a drastic intergroup surge among individuals aged 46 and above compared to age less than 45 over the study years. For instance, in 2020, the incidence increased from 0.05 % in the 31-45 age group to 0.16% in the 46-60 age group, which is 3.2 times higher. This surge was most pronounced in 2016 (3.25 times) and lowest in 2017 (2.6 times). Despite an increase in the 61+ age group compared to the 46-60 age group; the increase is less pronounced than the 31-45 to 46-60 age groups. The 61+ age group’s incidence rate was 1.27-1.46 times higher than that of the 46-60 age group over the study years.

**Figure 2.**
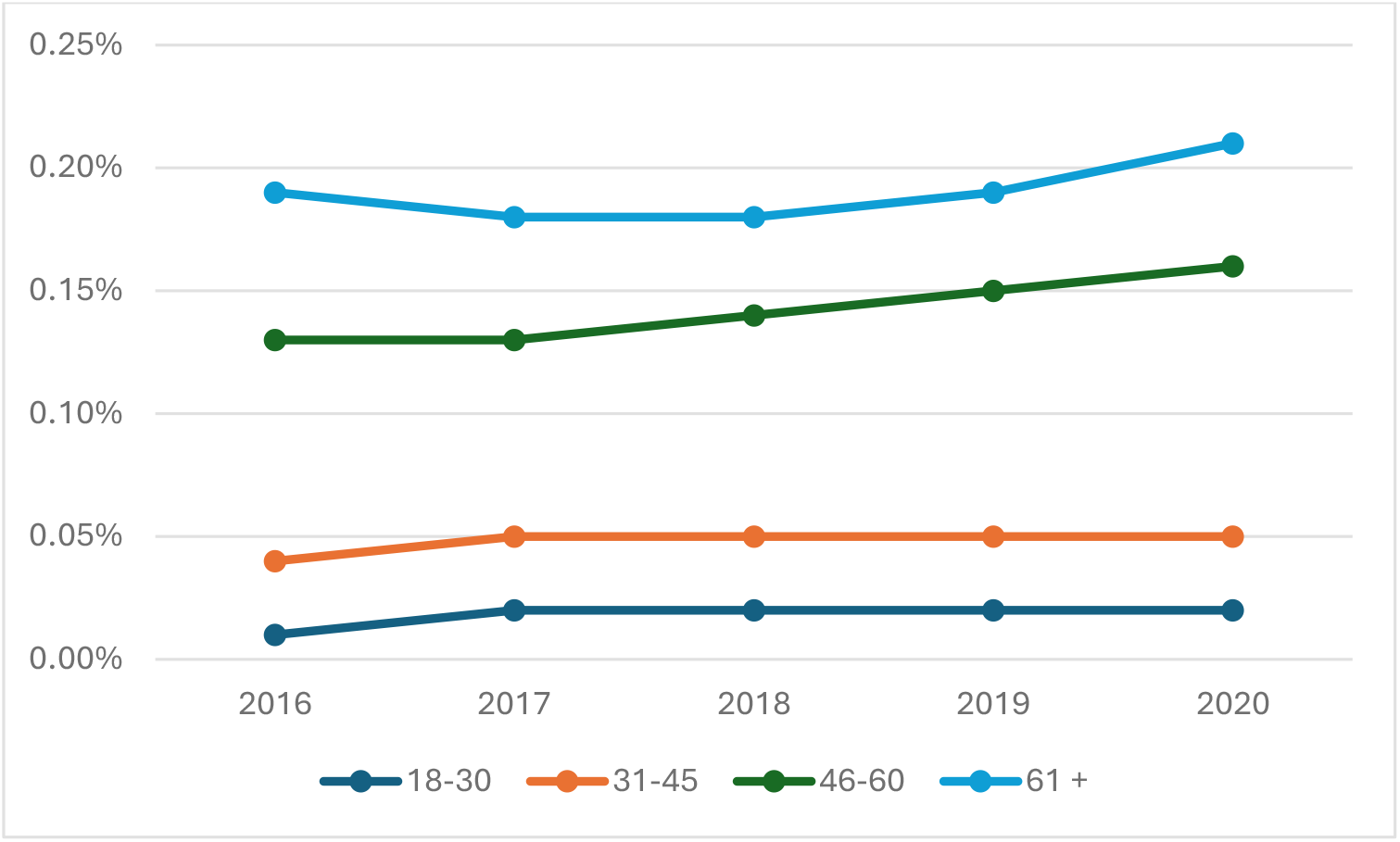
Takotsubo Cardiomyopathy Overall Incidence (2016-2020)

The data revealed noticeable racial disparities in incidence rates from 2016 to 2020 (Figure 3). White patients represented the largest portion of the study cohort, accounting for 0.16% of their population followed by native Americans at 0.13%, while the lowest incidence was among the black population (0.07%). Overall, the white population showed a continuous pronounced increase from 0.02% in 2016 to 0.18% in 2020. However, the native American population showed a decline, falling from 0.14% in 2016 to 0.11% in 2020. The Black population remained around 0.07%, with the slight increase to 0.08% in 2020. Incidence in other racial groups including Asians and Hispanics had similar behavior. Asian population revealed a gradual rise from 0.09 % to 0.13 % and Hispanic from 0.06 to 0.09%, from 2016 to 2020, respectively.

**Figure 3.**
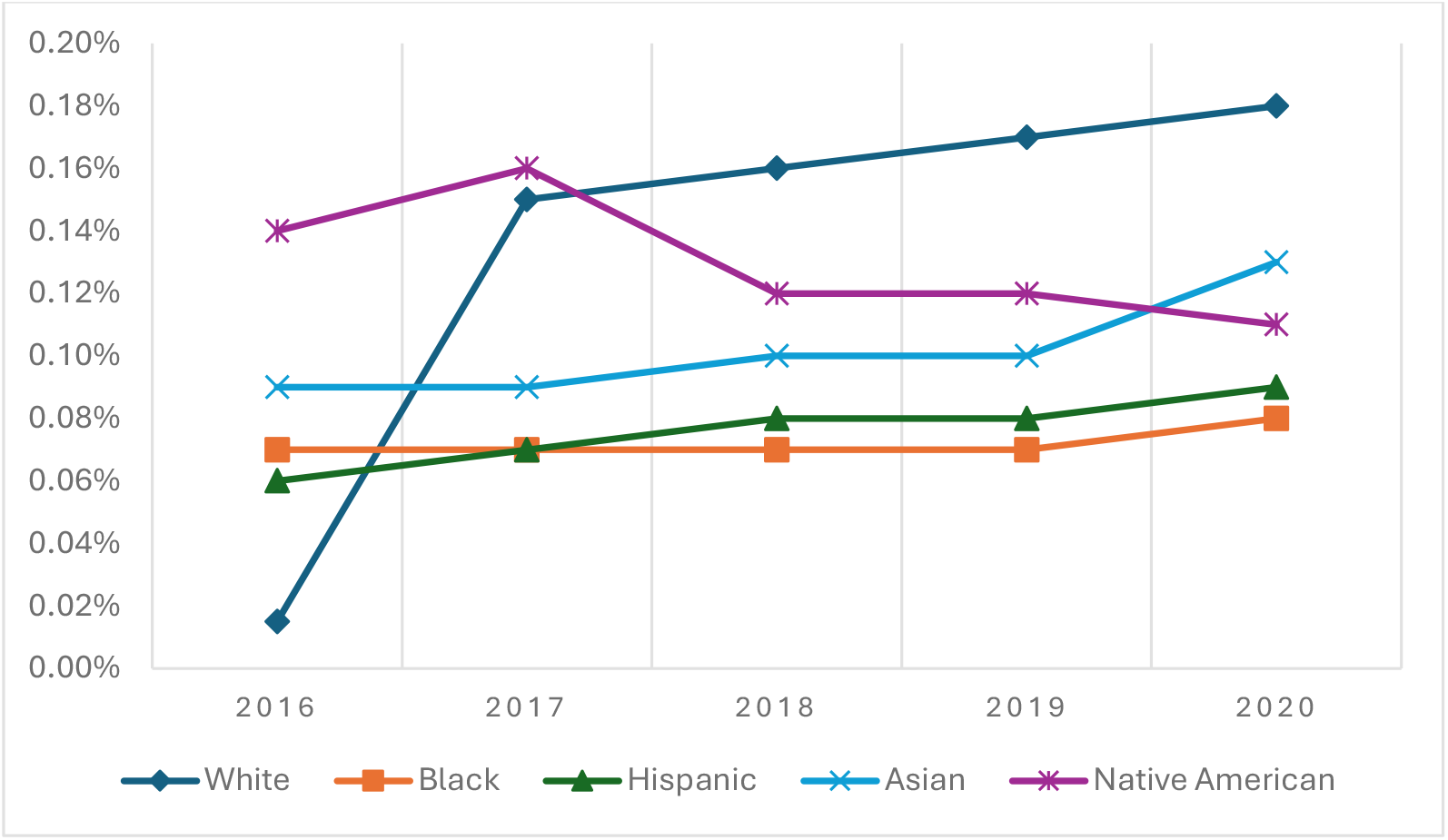
Takotsubo Cardiomyopathy Incidence Over Years Based on Race.

Socioeconomic factors including median household income, hospital bed size, and primary payer varied significantly. Patients with TC tend to have higher household incomes and larger hospital bed sizes. Most patients were on Medicare (0.18% vs 0.08% for Medicaid or no charge).

Takotsubo cardiomyopathy patients are admitted more in the private, non-profit, and urban teaching hospitals with the lowest admission percentage in southern areas.

### In-hospital complications

Patients with Takotsubo cardiomyopathy had a high burden of in-hospital mortality (6.58%) and cardiovascular complications including myocardial rupture (0.02%), cardiogenic shock (6.66%), atrial fibrillation (20.79%), cardiac arrest (3.42%), congestive heart failure (35.93%), and stroke (5.38%) compared to all patients. The odds ratio for in-hospital complications is shown in Table 2.

**Table 2.** In-Hospital Complications Total Non-Takotsubo Takotsubo P-value Odds Ratio.

| Table 2. In-Hospital Complications | Total | Non-Takotsubo | Takotsubo | P-value | Odds Ratio<br>(C.I.) |
| --- | --- | --- | --- | --- | --- |
| Mortality | 2.41% | 2.40% | 6.58% | <0.001 | 2.86(2.73-2.99) |
| Myocardial Rupture | 0.003% | 0.003% | 0.02% | <0.001 | 5.79(2.74-12.20) |
| Cardiogenic Shock | 0.57% | 0.56% | 6.66% | <0.001 | 12.71(12.19-13.26) |
| Atrial Fibrillation | 15.49% | 15.48% | 20.79% | <0.001 | 1.43(1.40-1.47) |
| Cardiac Arrest | 0.74% | 0.74% | 3.42% | <0.001 | 4.79(4.53-5.06) |
| Congestive Heart Failure | 13.77% | 13.74% | 35.93% | <0.001 | 3.52(3.44-3.60) |
| Stroke | 2.77% | 2.76% | 5.38% | <0.001 | 2.00(1.90-2.10) |

Overall mortality resulting from TC was significantly high at 6.58%, compared to 2.41% in all patients, with an odds ratio (OR) of 2.86 (95% CI, 2.73-2.99; P < 0.001), indicating 2.86 times higher risk of mortality compared to others. Patients with Takotsubo cardiomyopathy showed significantly higher odds of cardiogenic shock (OR 12.71; 95% CI, 12.19-13.26; P < 0.001), cardiac arrest (OR 4.79; 95% CI, 4.79(4.53-5.06); P < .001), congestive heart failure (OR 3.52; 95% CI, 3.44-3.60; P < .001), and myocardial rupture (OR 5.79; 95% CI, 2.74-12.20; P <0.001) as presented in Figure 4. In addition, patients with Takotsubo cardiomyopathy were 2 times more likely to experience a stroke (OR 2.00; 95% CI, 1.90-2.10; P < 0.001), and had a slightly higher incidence of atrial fibrillation (OR 1.43; 95% CI,1.40-1.47; P < 0.001).

**Figure 4.**
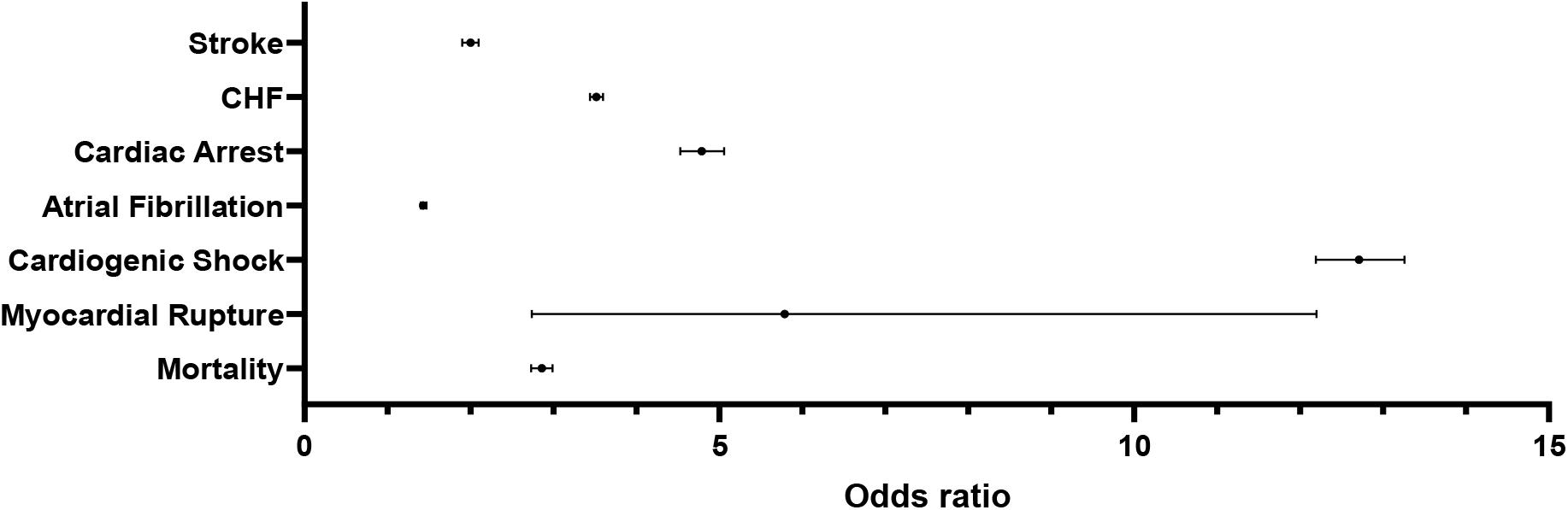
Forest Plot Showing Complications Related to Takotsubo Cardiomyopathy.

Over the years of the study, certain complications such as mortality, cardiogenic shock, cardiac arrest, congestive heart failure, and stroke have shown an apparent increase, while myocardial rupture and atrial fibrillation incidence remained relatively stable. Mortality increased from 5.63% to 8.38%, cardiogenic shock from 5.81% to 7.41%, cardiac arrest from 2.64% to 4.47%, CHF from 34.77% to 37.61%, and stroke from 4.97% to 5.9%, from 2016 to 2020, respectively.

Atrial fibrillation steady course escalated between 21.04% in 2016 to 21.31% in 2020. Myocardial rupture rate was very low, ranging between 0.00% to 0.03%. The most common complication was congestive heart failure followed by atrial fibrillation.

### Length of stay (LOS) and average total charges

Average LOS was constant over the years with a mean of 7(±10). However, the average of total charges increased constantly with the highest amount recorded in 2020. Based on multivariable linear regression, age, gender, race, median household income, hospital bed size, hospital region, primary payer, and hospital ownership/control type are all significantly associated with LOS (p<.001).

## Discussion

Our study of 199,920 patients hospitalized with Takotsubo cardiomyopathy in a large inpatient database from 2016 to 2020 revealed major insights into the epidemiology and complications of this condition over the years.

Our study discovered a new age-related pattern in patients with TC, identifying a critical turning point. It has been previously found that TC incidence increases with aging and usually follows a stressful situation, either emotional or physical [6]. Our study revealed a sudden increase in incidence in the 46-60 age group compared to 31-45 age group, with the older age group showing 2.6-3.25 times higher incidence. This sudden rise could be due to a combination of increased stress levels, hormonal variations, and the onset of cardiovascular risk factors along with under-treatment for hypertension and hyperlipidemia, alcohol use, and smoking [7-9].

Although our data did not differentiate between 4 various types of TC [10] due to the use of a single ICD-10 code, the recently diagnosed type of TC known as reverse-type Takotsubo cardiomyopathy tends to occur mainly in younger individuals [10]. The higher percentage of men in the current study might also contribute to this new age cut-off, as men tend to present with TC at a younger age than women [3, 11]. This new age-related finding holds important clinical implications. It could serve as a useful tool in discriminating between acute coronary syndrome and TC, aiding in the early diagnosis of this condition without a previous assumption that TC is an elderly-specific condition.

Consistent with previous studies [8], female gender was more likely to present with TC. The reason for this predominance is not fully understood. Hormonal changes especially in older ages might play a role [12, 13]. In a study by Ueyama et al, estradiol decreased the pathological changes in heart rate caused by emotional stress in rats [14]. However, the percentage of females in our study was lower (83%) compared with as high of 90% [10, 12], 97% [15], and 89% [16] in previous studies. This change might reflect previous underdiagnosis TC in men [1] and increased awareness of the condition in men.

Our observation of race showed that the lowest hospitalization for TC was among Caucasians in 2016 and the highest in 2020 among Caucasians followed by Native Americans. compatible with our results, in the study by Brinjikji et al, the majority of patients with TC were white [16]. In contrast to our results, Native Americans had the lowest frequency of TC using NIS data in 2008 [17]. Consistent with our results, Caucasian hospitalization increased over 2006-2014 in a nationwide sample study [4]. This finding might be related to better recognition of TC across all races, especially Caucasians.

Our results are compatible with prior studies in the USA regarding increased TC-associated hospitalization [4, 18]. It remains unclear whether the rise happened in the actual incidence, the number of diagnoses due to greater clinician awareness of the condition [19], or more frequent use of different diagnostic criteria, such as Mayo Clinic criteria [20]. Despite the studies on TC since its discovery in 1991 by Dote et al. [21], this condition is still not well understood from many perspectives. While catecholamine-induced myocardial injury is the most accepted mechanism for TC, the exact pathophysiology remains unknown [12]. Therefore, due to a completely poor understanding of pathophysiology, despite rigorous advancements in cardiac care, the treatment options in the acute TC phase are not completely determined and mortality of TC is still high. Mortality rates have been estimated at 0-8% in different studies [1], showing an incremental pattern over the years. Brinjikji et al. reported an in-hospital mortality rate of 4.2% for TC patients from 2008-2009 [16]. In a review by Singh et al. included 37 research papers from different regions, and overall in-hospital mortality was estimated at 4.5% [22]. In the Mayo Clinic Takotsubo syndrome registry, in-hospital mortality was 3% [23]. An interesting finding revealed by our study was that not only did mortality rates increase over the years of study, but also it was as high as 8.3% in 2020. This more than 1.5% increase in mortality from 2019 to 2020 can be attributed to multiple factors. Physical stress associated with TC is becoming more focus of attention and there are currently more studies investigating its incidence and prognosis [24]. The mortality rate resulting from TC related to physical stressors is higher [24, 25]. One of the most important recently discussed physical stressors is COVID-19 [26] . Covid 19 emerged in late 2019 and multiple studies reported significant increases in TC due to this pandemic [27, 28]. In the review by Chang et al, the inpatient mortality of patients with COVID and TC is higher than TC alone [29]. So, it seems that COVID might play a role in increasing the incidence and mortality rate in recent years [28].

Patients hospitalized with TC had substantial rates of other cardiovascular complications. Complications increased over the years, except for AF and myocardial rupture, which had a steadier course. Commensurate with other studies, congestive heart failure was the most common complication of TC [16] followed by atrial fibrillation. In a study by Templin et al, in hospital complications and death were similar to patients with acute coronary syndrome [30]. In another study by Vallabhajosyula et al., patients with TC had a higher risk of heart failure and lower risk of mortality compared to myocardial infarction during the 8 year period from 2007-2014 [31].

These data confirm that complications following TC are still high and even have incremental patterns over the years, mandating more research and intervention in this field.

### Strengths of This Study

There are several strengths to our study. The present study is one of the largest studies on TC assessing race, gender, and age differences along with a wide variety of complications and in-hospital mortality in a 5-year study course. Using nationally representative data allows the generalizability of the findings and eliminates the biases associated with small sample sizes and single-center studies.

### Limitations

The data was obtained from NIS using ICD-10 codes. As a result, they are prone to coding errors. We did not have data on different types of TC and the incidence of each subtype, hence associated outcomes with each type cannot be evaluated. NIS is an inpatient data register and did not have access to outpatient data, therefore long-term mortality cannot be evaluated. Another limitation is the lack of data on patients’ comorbid conditions which might contribute to their mortality. We did not have data on gender-specific mortality rates. Additional studies are warranted to mitigate these limitations.

## Conclusion

The data clearly indicate that individuals with Takotsubo syndrome are at significantly higher risk for a range of serious cardiovascular complications. Furthermore, there has been no improvement in any of the complications of interest throughout the years of the study. Further research on management and improvement in the care of these patients is warranted.

## Data Availability

Publically available

